# Peripheral blood immune status at clinical onset correlates with severity of Coronavirus Disease 2019

**DOI:** 10.1101/2020.06.03.20116533

**Authors:** Nathalie Santoro, Alessandro Cignetti, Valerio Tenace, Ines Clotilde Casonato, Domenico Cosseddu, Margherita Vizzini, Giovanni De Rosa, Massimo Geuna

## Abstract

The Coronavirus Disease 2019 (COVID-19) pandemic is a global threat to healthcare systems, requiring hospitalization in sub intensive and intensive care for respiratory syndrome in 25-30% of patients and accounting for a lethality up to 15%.

In this retrospective study the clinical characteristics of 215 COVID-19 patients were correlated with the peripheral blood immune status.

Different groups of COVID-19 patients may be identified on the basis of clinical behavior and a strong correlation between groups and age and comorbidities as well as with the immune profile is demonstrated. A lower age correlates with a lower severity of the disease differentiating between patients who may be quarantined at home and those requiring hospitalization. An older age (>82 years) together with a higher number of comorbidities is associated to a very severe prognosis. The absolute number of CD3+, CD4+, CD8+ T lymphocytes was progressively decreasing according to the severity of the disease and a CD3+ and CD8+ threshold indicating very severe cases is suggested.

## Introduction

The novel severe acute respiratory syndrome coronavirus 2 (SARS-CoV-2) outbreak has affected million people worldwide so far with over 300,000 confirmed death (1). The Coronavirus disease 2019 (COVID-19) presents with a highly heterogeneous spectrum of symptoms spanning from asymptomatic, to mild respiratory ones (fever, cough, shortness of breath), to severe (interstitial pneumonitis requiring intensive care and external respiratory support) up to death (2). The main predictive parameters associated with a greater severity of SARS-CoV-2 infection and with the clinical course of symptomatic and hospitalized COVID-19 patients have been identified in older age and in the presence of comorbidities (mainly diabetes mellitus, hypertension, cardiovascular diseases, chronic respiratory diseases) (2). Current available information on innate immune status of SARS-CoV-2-infected patients is the increased neutrophils counts, the reduced lymphocytes counts and the increased serum levels of IL-6 and of C-reactive protein, suggesting a strong inflammatory response (3). A first report on changes in lymphocyte populations in patients severely affected by Covid-19 indicate a low T cells count, an increase in naïve helper T cells and a decrease in memory helper T cells (4). The reduction in absolute count of peripheral T cells has also been associated to the severity of the disease (5, 6). The aim of the present study was to analyze the immune profile of peripheral blood lymphocytes in patients with COVID-19 at hospital admission and evaluate the correlation between the immune status and the other clinical and laboratory parameters.

## Materials and Methods

All patients (n=230) were admitted at emergency department with respiratory symptoms compatible with SARS-CoV-2 infection. All patients authorized data disclosure by signing an informed consent form at admission. All procedures performed in the study were in accordance with the ethical standards of the Helsinki Declaration.

Throat swab samples were obtained from patients and processed with RT-PCR method by means of the diagnostic GeneFinder COVID-19 PLUS RealAmp kit (OSANG Healtcare Co., Ltd, Korea), following manufacturer instructions, to confirm COVID-19 diagnosis.

The complete blood count with automated differential count was performed using Siemens Advia2120 hematology analyzer.

The peripheral blood (PB) immunophenotype was performed using a lyse no wash technique with single platform absolute count (Flow Count beads, Beckman Coulter, Milan, Italy). Briefly, 50 μl of peripheral blood, collected in EDTA containing tubes, were stained with a mixture of monoclonal antibodies (CD45 KO, HLA-DR PB, CD8 FITC, CD16+CD56 PE, CD19 PC7, CD4 APC, CD3 APC-AF750, all from Beckman Coulter). After 15 minutes of incubation in the dark, at room temperature (RT) erythrocyte were lysed with 1 ml of ammonium chloride solution for 15 minutes at RT, then 50 μl of paraformaldehyde solution were added, incubated 5 minutes and, immediately prior to cytometer analysis, 50 μl of Flow Count beads were added.

Samples were acquired with a 3-lasers 10-colors Navios cytometer and Navios software (Beckman Couter) collecting at least 500,000 total events or 10,000 beads. Analysis of the lymphocyte subpopulations was performed using a sequential gate strategy: doublet exclusion, debris/unlysed erythrocytes exclusion, CD45/SSC lymphocyte identification.

Bronchoalveolar lavage (BAL) liquid was filtered on a sterile gauze, the cells counted in Burker hemocytometer, then centrifuged 10 minutes at 400g, the supernatant was aspired, and the cell concentration adjusted at 5-10 × 10^6^ cells/ml in phosphate buffered solution (PBS). An amount of the cell suspension was cytocentrifuged, the slides dried and then stained with Diff-Quick staining for differential cell count. Hundred μl of the cell suspension were stained with the same mixture of antibodies used for PB, lysed with 2 ml of ammonium chloride solution, centrifuged 5 minutes at 400g, washed once with PBS, resuspended in 0,5 ml of PBS and immediately acquired to the cytometer.

The XLstat software was used for the statistical analysis (ANOVA, T test), data modelling and graphical representation (box and whiskers plots).

## Results

As of April 17^th^, 2020 a total of 230 patients have been admitted to the emergency department with respiratory symptoms compatible with SARS-CoV-2 infection. In 203 (88.3%) the diagnosis of covid-19 was confirmed by use of quantitative RT-PCR of throat swab samples. In 10 (4.3%) patients the SARS-CoV-2 infection was confirmed after a second or third throat swab sample. In two patients (0,9%) with 3 negative throat swab samples, but with clinical and radiological features compatible with COVID-19, the final diagnosis was obtained by use of QRT-PCR on bronchoalveolar lavage sample. In other 15 (6.5%) patients the diagnosis of COVID-19 was not confirmed and other causes of pneumonia were identified.

The data obtained from the retrospective analysis of 215 COVID-19 patients are summarized in Table 1. The ratio female/male was 1/1.9 and the mean age was 65.6 yrs, the average number of comorbidities was 1.7, ranging from 0 to 8. The most common comorbidities were diabetes mellitus, hypertension, cardiovascular diseases and neoplasia (data not shown). In all patients differential count of peripheral blood leucocytes and immunophenotype of peripheral blood lymphocytes were performed with absolute and relative count of CD3+, CD4+, CD8+, CD19+, CD16+CD56+ and CD3+HLADR+ (activated T lymphocytes) cell subsets. The analysis of the cytograms obtained with the hematology analyzer did not show any significant alteration in lymphocyte volume or shape compared to normal samples, as already described by Chong et al (7).

**Table 1.** Characteristic and laboratory data of 215 COVID-19 patients

|  | All patients (n=215) | Normal range |
| --- | --- | --- |
| Sex, F/M | 74/141 |  |
| Age, yrs, mean (range) | 65,6 (29-94) |  |
| Any comorbidity, mean (range) | 1,7 (0-8) |  |
| Treatment |  |  |
| Hydroxychloroquine, n (%) | 197 (91.6) |  |
| Antiviral drug, n (%) | 189 (97.9) |  |
| Tocilizumab, n (%) | 59 (27.4) |  |
| Lymphocyte count, % mean (range) | 19.7 (1.0-57.5) |  |
| x10<sup>9</sup>/L (range) | 1.00 (0.12-3.3) | 0.90-3.80 |
| Neutrophils count, % mean (range) | 74.7 (34.0-97.0) |  |
| x10<sup>9</sup>/L (range) | 5.07 (0.85-94.99) | 1.90-8.00 |
| Monocyte count, % mean (range) | 5.8 (1.0-16.3) |  |
| x10<sup>9</sup>/L (range) | 0.33 (0.04-2.30) | 0.16-1.00 |
| T lymphocytes CD3+, % mean (range) | 69.2 (30.7-94.5) | 59.0-85.0 |
| x10<sup>6</sup>/L (range) | 710.6 (73.0-3051.0) | 852-2618 |
| T lymphocytes CD3+CD4+, % mean (range) | 45.1 (9.9-76.3) | 31.0-59.0 |
| x10<sup>6</sup>/L (range) | 455.1 (31.0-1728.0) | 491-1699 |
| T lymphocytes CD3+CD8+, % mean (range) | 22.6 (4.7-58.7) | 15.0-45.0 |
| x10<sup>6</sup>/L (range) | 240.8 (23.0-1909.0) | 244-1115 |
| Activated T lymphocytes CD3+HLADR+, % mean (range) | 8.4 (0.9-38.1) | 2.0-21.0 |
| x10<sup>6</sup>/L (range) | 85.5 (9.0-1238.0) | 34-518 |
| NK cells CD3-CD16+CD56+, % mean (range) | 17.4 (1.3-63.3) | 4.0-28.0 |
| x10<sup>6</sup>/L (range) | 162.0 (12.0-780.0) | 77-661 |
| B lymphocytes CD19+, % mean (range) | 12.7 (0.0-45.4) | 5.0-20.0 |
| x10<sup>6</sup>/L (range) | 119.8 (0.0-551.0) | 70-488 |
| Ratio CD4/CD8, mean (range) | 2.6 (0.4-15.3) | 0.4-3.0 |
| Neutrophils/Lymphocytes ratio, mean (range) | 7.6 (0.6-97.0) |  |

Patients were divided in five groups (Series): the first (S1, n=117, 54.4%) group consisted of patients with low severity symptoms who, after diagnosis and laboratory screening, were quarantined at home. The second group (S2, n=51, 23.7%) was represented by patients with mild symptoms who needed to be hospitalized for supportive care and non-invasive ventilation (sub intensive care). In the third group (S3, n=24, 11.2%) there were patients undergoing intensive/invasive treatment (mechanical ventilation using Bilevel Positive Airway Pressure – BiPAP – or tracheotomy). The fourth group (S4, n = 14, 6.5%) consisted of patients who died early and whose clinical conditions at hospital admission were critical, without fulfilling criteria for intensive treatment. The fifth group of patients (S5, n=9, 4.2%) included those who underwent intensive care and died. The follow up of the 5 groups was restricted to 30 days.

Analysis of variance by ANOVA (Table 2) showed that, overall, the five groups differed significantly to each other for all parameter investigated, except for the absolute value of monocytes, the number of activated T lymphocytes (CD3+HLA-DR+) and the CD4/CD8 ratio. The analysis of the differences between group pairs by T test (Table 3) revealed that age was significantly lower in the S1 group (60.6 years) than in the other four groups (70.6, 68.4, 82.6 and 69.2 years respectively for S2, S3, S4 and S5). Similarly, the average age in S2, S3 and S5 was significantly lower than in S4. By contrast, there was no age difference in S2 vs. S3, S2 vs. S5, and S3 vs. S5.

**Table 2.** Differences between the 5 groups for all parameters (ANOVA test). abs: absolute count. ns: not significant.

| Parameter | F-stat | p-value |
| --- | --- | --- |
| Age | 13.717764 | 0.000000 |
| Comorbidities | 5.770737 | 0.000200 |
| Lymphocytes (abs) | 6.759675 | 0.000039 |
| Neutrophils (abs) | 5.576141 | 0.000276 |
| Monocytes (abs) | 2.414343 | ns |
| CD3+ (abs) | 7.162376 | 0.000020 |
| CD4+ (abs) | 6.413701 | 0.000069 |
| CD8 (abs) | 3.821702 | 0.005077 |
| CD3+DR+ (abs) | 2.060414 | ns |
| CD16+56+ (abs) | 3.271648 | 0.012537 |
| CD19+ (abs) | 2.834195 | 0.025532 |
| Ratio CD4/CD8 | 1.233168 | ns |
| NLR (ratio) | 16.604154 | 0.000000 |

**Table 3.** Clinical and laboratory findings in the 5 groups of COVID-19 patients. Comorb: comorbidities. Lymph: lymphocytes. Neut: neutrophils. Mono: monocytes. ns: not significant.

|  |  | Age, mean<br>(range) | Comorb,<br>mean<br>(range) | Lymph,<br>mean<br>x10<sup>9</sup>/L (%) | Neut, mean<br>x10<sup>9</sup>/L (%) | Mono,<br>mean<br>x10<sup>9</sup>/L (%) | Ratio<br>CD4/CD8,<br>mean | Ratio Neut/<br>Lymph,<br>mean |
| --- | --- | --- | --- | --- | --- | --- | --- | --- |
|  | Series 1 (S1)<br>(n=117) | 60.6<br>(29-88) | 1.4<br>(0-7) | 1.10<br>(22.3) | 4.04<br>(71.7) | 0.31<br>(6.0) | 2.50 | 4.86 |
|  | Series 2 (S2)<br>(n=51) | 70.6<br>(29-90) | 2.2<br>(0-8) | 1.07<br>(21.7) | 4.74<br>(72.1) | 0.41<br>(6.6) | 2.73 | 5.97 |
|  | Series 3 (S3)<br>(n=24) | 68.4<br>(50-80) | 1.4<br>(0-4) | 0.58<br>(10.1) | 6.45<br>(86.3) | 0.32<br>(4.8) | 2.78 | 15.70 |
|  | Series 4 (S4)<br>(n=14) | 82.6<br>(68-94) | 2.9<br>(1-6) | 0.83<br>(13.1) | 12.35<br>(82.9) | 0.38<br>(3.8) | 2.24 | 13.69 |
|  | Series 5 (S5)<br>(n=9) | 69.2<br>(55-84) | 1.8<br>(1-4) | 0.58<br>(9.4) | 6.82<br>(88.1) | 0.19<br>(2.5) | 3.80 | 22.50 |
| Test T,<br>p value | S1 vs S2 | 1.26E-05 | 1.29E-03 | ns | ns | 1.59E-02 | ns | ns |
|  | S1 vs S3 | 1.92E-04 | ns | 4.97E-09 | 5.51E-04 | ns | ns | 4.01E-04 |
|  | S1 vs S4 | 4.02E-10 | 3.20E-03 | ns | ns | ns | ns | 1.75E-02 |
|  | S1 vs S5 | 1.42E-02 | ns | 1.83E-03 | 9.19E-03 | 4.02E-03 | ns | ns |
|  | S2 vs S3 | ns | 4.19E-03 | 1.12E-06 | 1.90E-02 | ns | ns | 1.31E-03 |
|  | S2 vs S4 | 3.95E-05 | ns | ns | ns | ns | ns | 3.15E-02 |
|  | S2 vs S5 | ns | ns | 2.88E-03 | 3.65E-02 | 1.46E-04 | ns | ns |
|  | S3 vs S4 | 2.50E-06 | 3.37E-03 | ns | ns | ns | ns | ns |
|  | S3 vs S5 | ns | ns | ns | ns | 3.87E-03 | ns | ns |
|  | S4 vs S5 | 1.45E-03 | 3.20E-02 | ns | ns | ns | 4.66E-02 | ns |

The average number of comorbidities was significantly different in S1 (1.4) vs. S2 (2.2), S1 vs S4 (2.9), S2 vs S3 (1.4), S3 vs S4 and S4 vs S5 (1.8). Taken together, the most significant difference between groups was the older age and the higher degree of comorbidities observed in group 4.

The lymphocyte number was significantly lower in the S3 and S5 group compared to both S1 and S2, while, comparing the very same groups, the granulocyte number was higher. These data suggest that patients undergoing intensive care (i.e. patients with severe disease) display higher granulocyte and lower lymphocyte counts than group 1 and 2, (i.e. patients with milder disease not requiring intensive care). Patients in group 4, having a shorter course of the disease, behave differently from all other patients, displaying high neutrophil counts and moderate lymphopenia. The neutrophil/lymphocyte ratio (NLR) reflected this pattern for the most, being significantly different in S3 vs S1 and S2, S4 vs S1 and S2, while the difference of S5 vs S1 and S2, despite of the very different mean values, did not reach significance, probably because of the small number of patients in S5 (Table 3). S5 was the group that accounted for the most striking differences in monocyte counts, being significantly lower when comparing S5 vs S3, S2 and S1. Monocytes were also reduced in S2 compared to S1.

Mean value, standard deviation and range of CD3+, CD4+, CD8+, CD3+HLADR+, CD19+ and NK cells for each different group was compared and is illustrated in figure 1. There were no significant differences between the absolute number of each lymphocyte subset of the S1 group compared to S2 and S4 (panel A and C). By contrast, almost all lymphocyte subsets were significantly reduced in S3 and S5 groups compared to the S1 (panel B and D). Similarly, the absolute number of CD3+, CD4+, CD8+, CD3+HLADR+ and NK cells was reduced in S3 compared to S2 (panel E). Finally, a significant reduction of CD3+ and CD8+ lymphocytes was observed in S5 compared to S4 (panel F). Notably, the number of CD8+ cells is 97−10^6^ /L in S5 and 211−10^6^ /L in S4. These data suggest that a low absolute number of CD3+ cells accounts for the lower lymphocyte counts observed in patients with severe disease compared to patients with milder disease.

**Figure 1.**
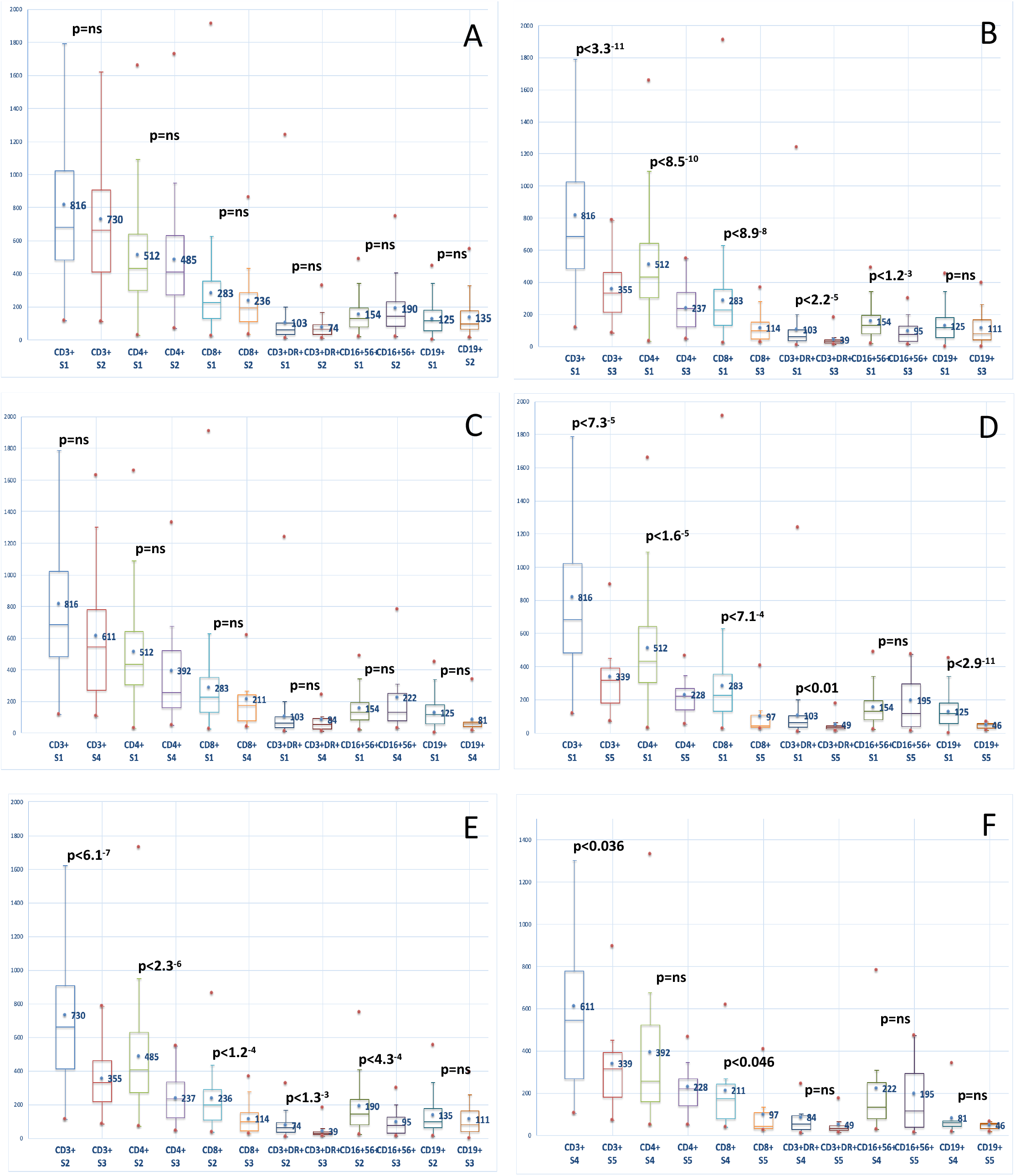
Graphic representation (box and whiskers) of the distribution of peripheral blood lymphocyte populations (value in cells/106/L) in the different groups (S1, S2, S3, S4, S5) of COVID-19 patients. The number reported in each box is the mean value of cells for each cell population in each patient group. A, group S1 vs S2. B, group S1 vs S3. C group S1 vs S4. D, group S1 vs S5. E, group S2 vs S3. F, group S4 vs S5. P value is referred to T test between the two groups. Analysis and graphs obtained with XLSTAT software.

In two patients, after three negative results for detection of SARS-CoV-2 RNA by QRT-PCR on swab throat samples, a bronchoalveolar lavage (BAL) with diagnostic purpose was performed and resulted positive. In both BAL a differential count on cytocentrifuged cells and a flow cytometry lymphocyte analysis were obtained. In both samples an increased percentage of lymphocytes (63% and 60% respectively) compared to normal values (10-15%) (8) was observed, with an increased percentage of CD8 T-cells (Figure 2).

**Figure 2.**
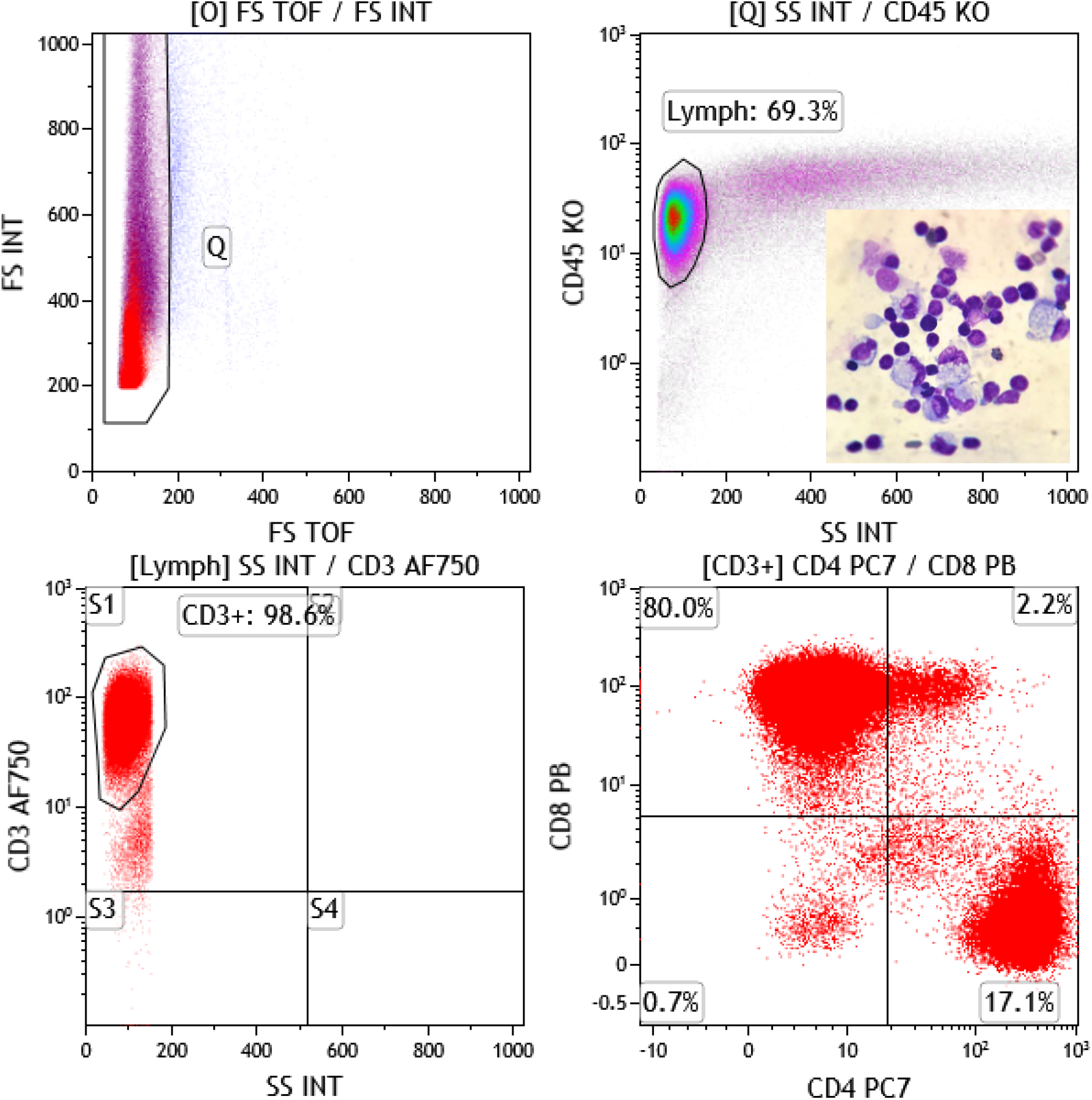
Flow cytometry of BAL sample. The first plot (FS TOF/FS INT) shows the gate for doublet exclusion, the second (SS/CD45) t he lymphocytes g ate, and the third (SS/CD3) the gate for T lymphocyte. In the fourth plot (CD4/CD8) the distribution of CD4 and CD8 T cells gated on CD3+ cells is shown. In the insert, the cell morphology of one representative BAL sample shows the high number of lymphocytes.

## Discussion

The pivotal role of T cell mediated immunity in viral infection and clearance is well known for CD4+ helper T cells in assisting and enhancing the function of cytotoxic T cell and B cell, and for CD8+ cytotoxic T cells in contributing directly to virus eradication by means of perforin and cytokines (9, 10). However, the persistence of viral stimulation may induce a shift in the differentiation pathway of CD4+ T cells and T cell exhaustion (9). Earlier studies in COVID-19 patients showed a reduction of T cells and an increase of exhausted T cells expressing PD-1 antigen, as well as a direct correlation between T cell reduction and severity of symptoms (11). In this retrospective analysis of 215 COVID-19 patients we correlate the immune status at hospitalization with clinical behavior, age and comorbidities, to explore if some immunological parameter may be predictive of the clinical course. As previously reported (2), also in our series we observed a low female/male ratio, an average age greater than 65 years and a significant presence of comorbidities. The correlation between severity of symptoms, age and number of comorbidities is clear between the different groups of patients with the exception of that between groups S2 and S3, S2 and S5, S3 and S5 by age and between groups S1 and S3, S1 and S5, S2 and S4 and S5, S3 and S5 for the number of comorbidities.

These data seem to indicate that age is not a discriminating factor among patients who require hospitalization. Conversely, the number of comorbidities is greater in patients who die without intensive care (S4) than in patients undergoing intensive care (S3 and S5). The immune status, in term of absolute number of cells, is more compromised in patients who need intensive care compared to other patients.

This is true not only comparing patients with low to mild severity symptoms (S1) with patients in sub intensive (S2) or intensive (S3) therapy, but also comparing sub-intensive (S2) with intensive care (S3) patients. Finally, despite the small number of patients in each group, a significant difference between S4 and S5 is present regarding age (lower in S5), comorbidities (lower in S5), absolute value of CD3+ and CD8+ (extremely reduced in S5). Taken together, these data seem to indicate that a reduction in lymphocytes number, particularly CD3+ and CD8+, is predictive of a more severe clinical course. The differences observed in the groups S4 and S5 could suggest that in the S4 group COVID-19 disease may be a concurrent cause of death (older patients, many comorbidity) while in group S5 COVID-19 disease, of which deep immune depression represents one of the hallmarks, can be considered the main cause of death. Similarly, in S2 and S3 the immune system results even more involved in discriminating patients with different severity of symptoms, being reduced in S3 also CD4+, NK and activated T cells. The reduction of T cells, mainly CD8 T lymphocytes, opens several questions on the fate of these cells during SARS-CoV-2 infection. One possible explanation is tissue recruitment and segregation of CD8+ T cells at infection site (i.e. lung) where they contribute to acute respiratory distress syndrome (ARDS). On support of this hypothesis we observed, in two bronchoalveolar lavages of COVID-19 patients, a predominant lymphocytic infiltration mainly sustained by CD8+ cells (Figure 2).

Further investigations on greater number of patients are needed to define a reliable threshold of CD3+ and CD8+ T cells below which the probability of a severe disease increases significantly (12). Nevertheless, we can estimate that in younger patients with no of few comorbidities, the absolute value of 339−10^6^ /L CD3+ and 97−10^6^ /L CD8+ (both values are the mean value of S5 group) could be scored as negative indicators.

The present study has some limitations. First of all, we only evaluated the main lymphocyte populations without investigating T cell subsets or cytokine production. Secondly, it is a retrospective study that does not take into account second or third immunophenotypic analysis of the same patients and the obvious evolution of the immune profile. Similarly, the follow up of the patients is limited to a short period of time, and consequently the group to which they are belonging reflects a “frozen” situation. On the other hand, the purpose of the present study was not to evaluate the clinical course of the disease but to identify one or more early indicators of the clinical course, focusing on clinical and laboratory features at diagnosis.

In conclusion, the immune profiling associated to age and comorbidities may help identifying different groups of COVID-19 patients with different clinical behavior. Further studies are needed to identify laboratory parameters correlated with the clinical course and to create a scoring system that will rapidly guide therapy and clinical management.

## Data Availability

The clinical data used to support the findings of this study are restricted in order to protect patient privacy. Clinical data are available from the corresponding author upon request for researchers who meet the criteria for access to confidential data.
The laboratory data used to support the findings of this study are available from the corresponding author upon request.

## Notes

### Competing Interest Statement

The authors have declared no competing interest.

### Clinical Trial

The study is a preliminary retrospective analysis in a setting of health emergency. The project of a clinical trial is under submission to the Ethics Committee.

### Funding Statement

The work presented did not receive external fund.

### Author Declarations

The IRB (Hospital Medical Direction) approval for the use of clinical and laboratory data for the present study has been obtained.

